# Safety Monitoring of Bivalent COVID-19 mRNA Vaccines Among Recipients 6 months and Older in the United States

**DOI:** 10.1101/2024.01.24.24301676

**Authors:** Patricia C. Lloyd, Elizabeth R. Smith, Joann F. Gruber, Michelle Ondari, Hui Lee Wong, Mao Hu, Tainya C. Clarke, Rowan McEvoy, Kandace L. Amend, Daniel C. Beachler, Cheryl N McMahill-Walraven, John D. Seeger, Alex Secora, Djeneba Audrey Djibo, Jennifer Song, Nandini Selvam, Jonathan P. DeShazo, Robin Clifford, Eugenio Abente, Yoganand Chillarige, Richard A. Forshee, Steven A. Anderson, Azadeh Shoaibi

## Abstract

Active monitoring of health outcomes after COVID-19 vaccination provides early detection of rare outcomes post-licensure.

**Objective:** To evaluate health outcomes following bivalent COVID-19 Pfizer-BioNTech (BNT162b2) and Moderna (mRNA-1273.222) vaccination among individuals 6 months and older in the United States.

**Design:** Monthly monitoring of health outcomes from August 2022 to July 2023 in four administrative claims databases. Descriptive analyses monitored vaccine uptake, outcome counts and coadministration of bivalent COVID-19 and influenza vaccines. Sequential analyses tested for elevated risk of each outcome in a prespecified post-vaccination risk interval, or a period of hypothesized elevation based on clinical guidance, compared to a historical baseline.

**Participants and Exposures:** Persons 6 months and older who received a bivalent COVID-19 BNT162b2 or mRNA-1273.222 vaccine during the study period, with continuous enrollment in a medical insurance plan from the start of an outcome-specific clean interval to the COVID-19 vaccination date. Vaccines were identified using product-specific codes from medical coding systems.

**Health Outcomes:** Twenty outcomes were monitored in BNT162b2 vaccine recipients 6 months-4 years, and mRNA-1273.222 vaccine recipients 6 months-5 years. Twenty-one outcomes were monitored in BNT162b2 vaccine recipients 5-17 years and mRNA-1273.222 vaccine recipients 6-17 years. Eighteen outcomes were monitored in persons 18 years and older for both mRNA vaccines.

**Results:** Overall, 13.9 million individuals 6 months and older received a single bivalent COVID-19 mRNA vaccine. The statistical threshold for a signal was met for two outcomes in one database: anaphylaxis following bivalent BNT162b2 and mRNA-1273.222 vaccines in persons 18-64 years and myocarditis/pericarditis following bivalent BNT162b2 vaccines in individuals 18-35 years. There were no signals identified in young children.

**Conclusions:** Results were consistent with prior observations from published studies on COVID-19 vaccine safety. This study supports the safety profile of bivalent COVID-19 mRNA vaccines and the conclusion that the benefits of vaccination outweigh the risks.

## 1. Background

The U.S. Food & Drug Administration (FDA) first authorized bivalent formulations of COVID-19 mRNA vaccines in August 2022.^1^ These updated vaccines included both the SARS-CoV-2 ancestral strain and omicron BA.4 and BA.5 subvariants and offered added protection against more recently circulating strains of the virus.^1^ Vaccines were first authorized in persons 12 years and older for the bivalent COVID-19 Pfizer-BioNTech (BNT162b2) vaccine, and 18 years and older for the bivalent COVID-19 Moderna (mRNA-1273.222) vaccine.^2,3^ The authorization for both vaccine brands was expanded in October 2022 to include persons 5 years and older for the bivalent BNT162b2 vaccine and 6 years and older for the mRNA-1273.222 vaccine.^2,3^ In December 2022, the authorization for both vaccine brands was further expanded to include populations 6 months and older.^2,3^

There have been approximately 56 million bivalent COVID-19 vaccine doses administered in the U.S. as of May 2023.^4^ Of these, 42.0% have been administered to persons 65 years and older, 51.9% to persons aged 18-64 years, 5.9% to persons aged 5-17 years, and <1% to the population aged 6 months-4 years.^4^ Overall, bivalent COVID-19 mRNA vaccines have been administered to about 17.0% of the U.S. population.^4^

FDA has been performing near real-time surveillance to monitor the real-world safety of bivalent COVID-19 vaccines available in the U.S. Near real-time surveillance is a screening method designed to rapidly identify potential safety signals as vaccines are administered. However, results do not establish a causal association due to methodologic limitations. More robust epidemiological studies may be used to evaluate potential safety signals identified from screening methods.

This study presents results from FDA’s near real-time safety monitoring of bivalent COVID-19 vaccines using data from three commercial databases representing vaccine recipients aged 6 months to 64 years, and the Centers for Medicare & Medicaid Services (CMS) Medicare database representing vaccine recipients aged 65 years and older.

## 2. Methods

### 2.1 Data Sources

This study used administrative commercial health claims data from Carelon Research, CVS Health, and Optum to capture health care data on the population aged 6 months – 64 years old. Medicare Fee-For-Service (FFS) data from the CMS Shared Systems Database was used to capture data on the population aged 65 years and older. These databases contain longitudinal medical and pharmacy claims data that captures patient’s demographic information, clinical diagnoses, and vaccine administration information among other health care utilization information. The CMS Medicare enrollment database was used to capture enrollment information for the Medicare population. Where available, local and state-based Immunization Information Systems (IIS) data was linked to commercial claims databases to supplement the capture of patient’s COVID-19 vaccination history. Once validated, data from select IIS jurisdictions was included in the analysis. Supplementary Table E1 summarizes individual database-related characteristics including enrollment size and claims delay.

### 2.2 Study Population and Period

Health plan members aged 6 months and older were included in surveillance if they received a bivalent COVID-19 mRNA vaccine during the vaccine brand- and age-specific authorization periods and were continuously enrolled in their respective health plan for the complete duration of the outcome-specific clean interval. The surveillance period extended from the U.S. authorization date for the bivalent COVID-19 mRNA vaccines (i.e., August 2022 for ages 12 years and older; October 2022 for 5-11 years of age; December 2022 for 6 months to 4 years of age) through mid-2023. Exact surveillance start dates varied by vaccine brand and age group based on vaccine authorization dates (Supplementary Table E2).

### 2.3 Exposures and Follow-Up

The exposure was defined as the receipt of the bivalent BNT162b2 or mRNA-1273.222 COVID-19 vaccines in any setting during the vaccine brand and age-specific authorization periods. Bivalent COVID-19 mRNA vaccines were identified in claims and IIS data using product and dose-specific codes from the Current Procedural Terminology (CPT)/ Health Care Common Procedure Coding System (HCPCS), National Drug Codes (NDCs), and CVX (vaccine administered) codes.^5^ Only individuals’ first bivalent vaccine administration was included in surveillance because bivalent COVID-19 vaccines were authorized as a single dose at the start of surveillance.

Follow-up time included all person-time accrued during the prespecified post-vaccination risk intervals. Risk intervals were defined as a period of hypothesized elevated risk due to vaccination and were established based on guidance from clinicians and literature review. Risk intervals were censored at disenrollment, subsequent COVID-19 vaccination, surveillance end date, or death.

### 2.4 Health Outcomes

Health outcomes were selected based on literature review, consultation with clinicians, and prior COVID-19 vaccine surveillance activities. Outcomes were monitored separately: an individual with multiple different outcomes was included in surveillance for each outcome, but only the first eligible occurrence of an outcome was included in the analysis. Eligible outcomes were defined as those occurring in the risk interval with no prior occurrence of the respective outcome during the outcome-specific clean interval. The clean interval was a period defined relative to vaccination date used to identify incident outcomes, in which an individual enters the outcome-specific study cohort only if the outcome did not occur during this interval. Table 1 presents the outcomes with clinical care settings, risk intervals, and clean intervals used to identify incident outcomes. It also indicates outcomes that were included in descriptive monitoring and sequential testing, by age group.

**Table 1.**
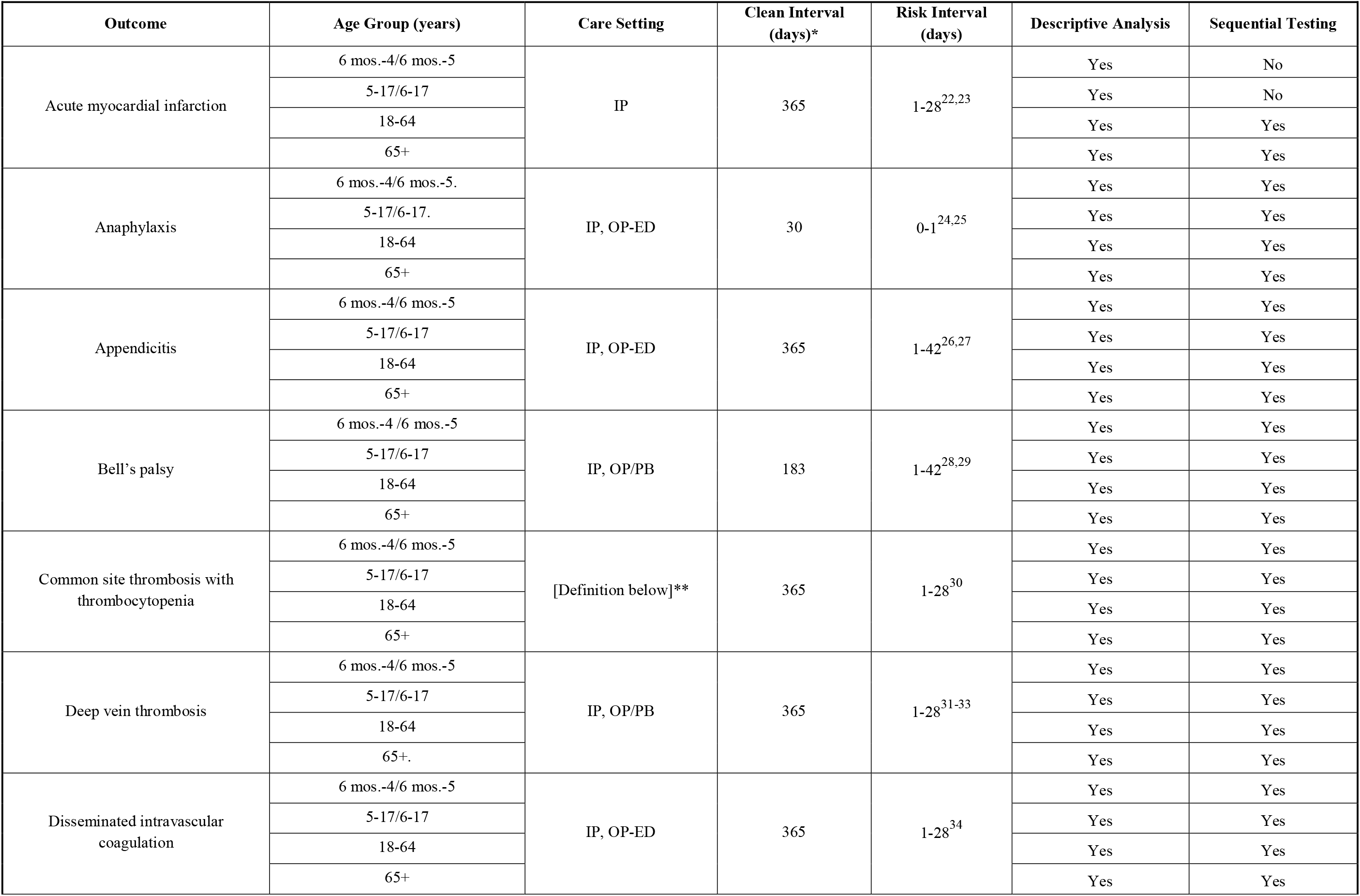
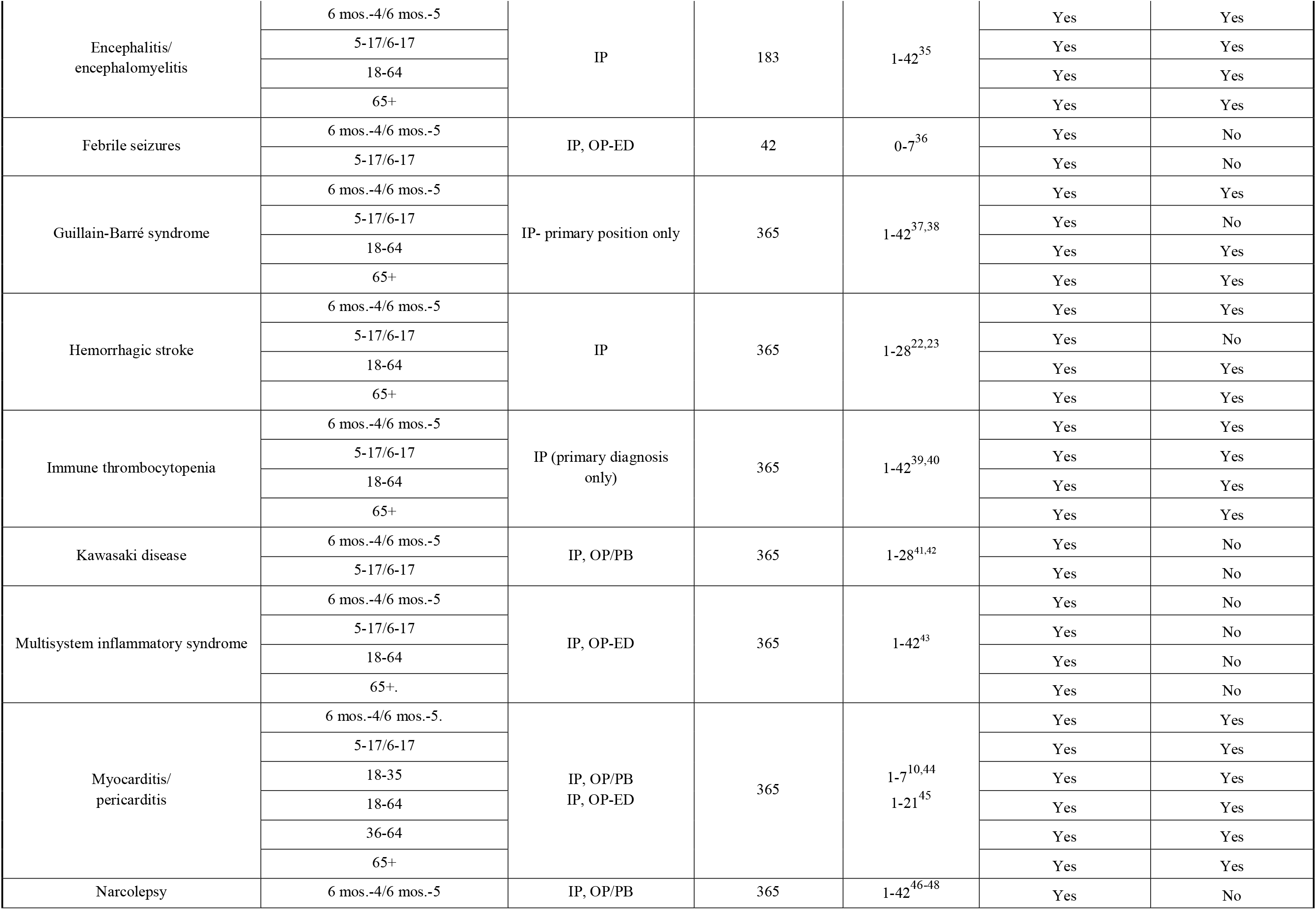
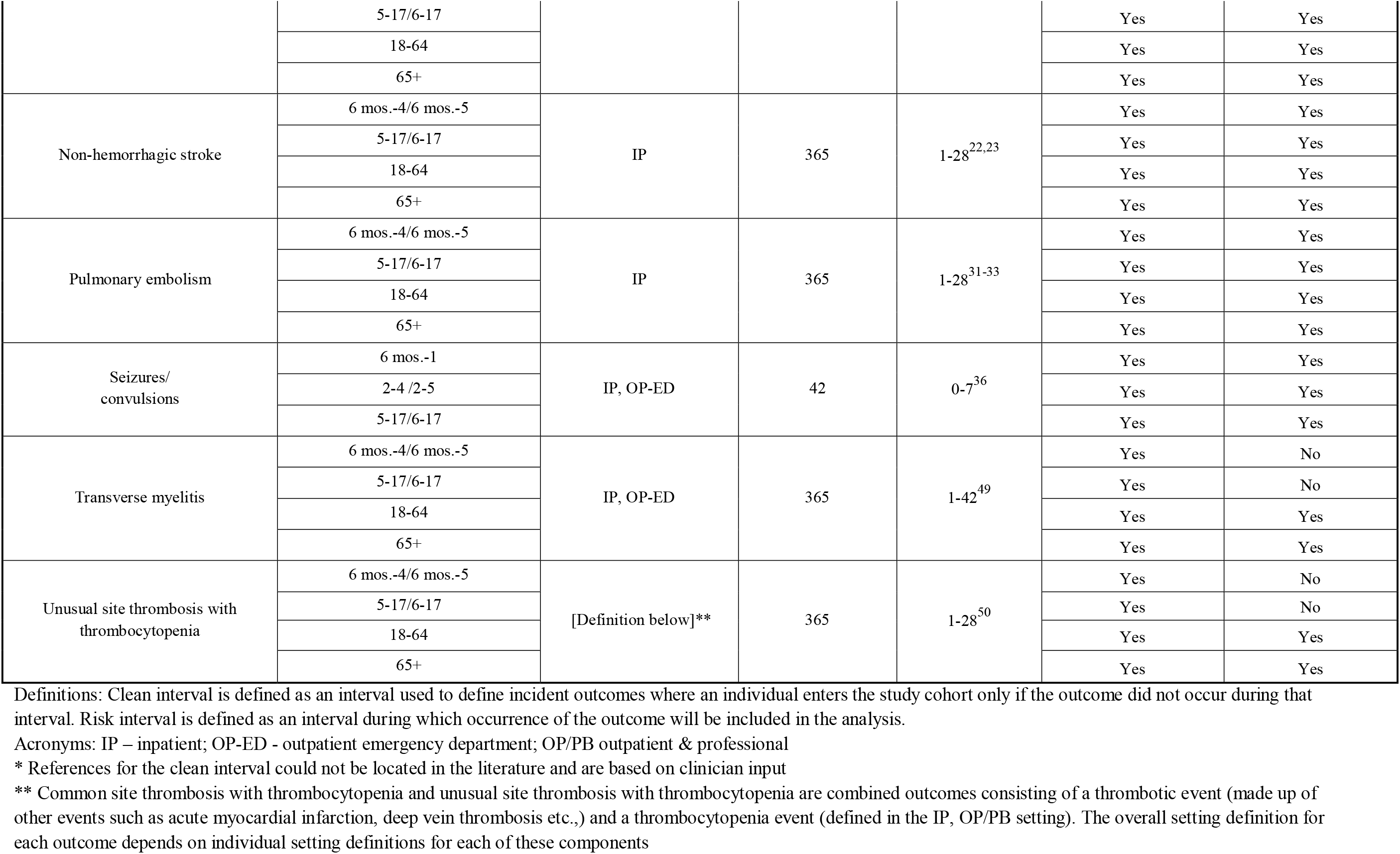
Health outcome definitions (settings, clean intervals, risk intervals), and analysis types.

| Outcome | Age Group (years) | Care Setting | Clean Interval (days)* | Risk Interval (days) | Descriptive Analysis | Sequential Testing |
| --- | --- | --- | --- | --- | --- | --- |
| Acute myocardial infarction | 6 mos.-4/6 mos.-5 | IP | 365 | 1-28<sup>22,23</sup> | Yes | No |
|  | 5-17/6-17 |  |  |  | Yes | No |
|  | 18-64 |  |  |  | Yes | Yes |
|  | 65+ |  |  |  | Yes | Yes |
| Anaphylaxis | 6 mos.-4/6 mos.-5. | IP, OP-ED | 30 | 0-1<sup>24,25</sup> | Yes | Yes |
|  | 5-17/6-17. |  |  |  | Yes | Yes |
|  | 18-64 |  |  |  | Yes | Yes |
|  | 65+ |  |  |  | Yes | Yes |
| Appendicitis | 6 mos.-4/6 mos.-5 | IP, OP-ED | 365 | 1-42<sup>26,27</sup> | Yes | Yes |
|  | 5-17/6-17 |  |  |  | Yes | Yes |
|  | 18-64 |  |  |  | Yes | Yes |
|  | 65+ |  |  |  | Yes | Yes |
| Bell's palsy | 6 mos.-4/6 mos.-5 | IP, OP/PB | 183 | 1-42<sup>28,29</sup> | Yes | Yes |
|  | 5-17/6-17 |  |  |  | Yes | Yes |
|  | 18-64 |  |  |  | Yes | Yes |
|  | 65+ |  |  |  | Yes | Yes |
| Common site thrombosis with thrombocytopenia | 6 mos.-4/6 mos.-5 | [Definition below]** | 365 | 1-28<sup>30</sup> | Yes | Yes |
|  | 5-17/6-17 |  |  |  | Yes | Yes |
|  | 18-64 |  |  |  | Yes | Yes |
|  | 65+ |  |  |  | Yes | Yes |
| Deep vein thrombosis | 6 mos.-4/6 mos.-5 | IP, OP/PB | 365 | 1-28<sup>31-33</sup> | Yes | Yes |
|  | 5-17/6-17 |  |  |  | Yes | Yes |
|  | 18-64 |  |  |  | Yes | Yes |
|  | 65+. |  |  |  | Yes | Yes |
| Disseminated intravascular coagulation | 6 mos.-4/6 mos.-5 | IP, OP-ED | 365 | 1-28<sup>34</sup> | Yes | Yes |
|  | 5-17/6-17 |  |  |  | Yes | Yes |
|  | 18-64 |  |  |  | Yes | Yes |
|  | 65+ |  |  |  | Yes | Yes |
| Encephalitis/<br>encephalomyelitis | 6 mos.-4/6 mos.-5 | IP | 183 | 1-42<sup>35</sup> | Yes | Yes |
|  | 5-17/6-17 |  |  |  | Yes | Yes |
|  | 18-64 |  |  |  | Yes | Yes |
|  | 65+ |  |  |  | Yes | Yes |
| Febrile seizures | 6 mos.-4/6 mos.-5 | IP, OP-ED | 42 | 0-7<sup>36</sup> | Yes | No |
|  | 5-17/6-17 |  |  |  | Yes | No |
| Guillain-Barré syndrome | 6 mos.-4/6 mos.-5 | IP- primary position only | 365 | 1-42<sup>37,38</sup> | Yes | Yes |
|  | 5-17/6-17 |  |  |  | Yes | No |
|  | 18-64 |  |  |  | Yes | Yes |
|  | 65+ |  |  |  | Yes | Yes |
| Hemorrhagic stroke | 6 mos.-4/6 mos.-5 | IP | 365 | 1-28<sup>22,23</sup> | Yes | Yes |
|  | 5-17/6-17 |  |  |  | Yes | No |
|  | 18-64 |  |  |  | Yes | Yes |
|  | 65+ |  |  |  | Yes | Yes |
| Immune thrombocytopenia | 6 mos.-4/6 mos.-5 | IP (primary diagnosis only) | 365 | 1-42<sup>39,40</sup> | Yes | Yes |
|  | 5-17/6-17 |  |  |  | Yes | Yes |
|  | 18-64 |  |  |  | Yes | Yes |
|  | 65+ |  |  |  | Yes | Yes |
| Kawasaki disease | 6 mos.-4/6 mos.-5 | IP, OP/PB | 365 | 1-28<sup>41,42</sup> | Yes | No |
|  | 5-17/6-17 |  |  |  | Yes | No |
| Multisystem inflammatory syndrome | 6 mos.-4/6 mos.-5 | IP, OP-ED | 365 | 1-42<sup>43</sup> | Yes | No |
|  | 5-17/6-17 |  |  |  | Yes | No |
|  | 18-64 |  |  |  | Yes | No |
|  | 65+. |  |  |  | Yes | No |
| Myocarditis/<br>pericarditis | 6 mos.-4/6 mos.-5. | IP, OP/PB<br>IP, OP-ED | 365 | 1-7<sup>10,44</sup><br>1-21<sup>45</sup> | Yes | Yes |
|  | 5-17/6-17 |  |  |  | Yes | Yes |
|  | 18-35 |  |  |  | Yes | Yes |
|  | 18-64 |  |  |  | Yes | Yes |
|  | 36-64 |  |  |  | Yes | Yes |
|  | 65+ |  |  |  | Yes | Yes |
| Narcolepsy | 6 mos.-4/6 mos.-5 | IP, OP/PB | 365 | 1-42<sup>46-48</sup> | Yes | No |
|  | 5-17/6-17 |  |  |  | Yes | Yes |
|  | 18-64 |  |  |  | Yes | Yes |
|  | 65+ |  |  |  | Yes | Yes |
| Non-hemorrhagic stroke | 6 mos.-4/6 mos.-5 | IP | 365 | 1-28<sup>22,23</sup> | Yes | Yes |
|  | 5-17/6-17 |  |  |  | Yes | Yes |
|  | 18-64 |  |  |  | Yes | Yes |
|  | 65+ |  |  |  | Yes | Yes |
| Pulmonary embolism | 6 mos.-4/6 mos.-5 | IP | 365 | 1-28<sup>31-33</sup> | Yes | Yes |
|  | 5-17/6-17 |  |  |  | Yes | Yes |
|  | 18-64 |  |  |  | Yes | Yes |
|  | 65+ |  |  |  | Yes | Yes |
| Seizures/<br>convulsions | 6 mos.-1 | IP, OP-ED | 42 | 0-7<sup>36</sup> | Yes | Yes |
|  | 2-4 /2-5 |  |  |  | Yes | Yes |
|  | 5-17/6-17 |  |  |  | Yes | Yes |
| Transverse myelitis | 6 mos.-4/6 mos.-5 | IP, OP-ED | 365 | 1-42<sup>49</sup> | Yes | No |
|  | 5-17/6-17 |  |  |  | Yes | No |
|  | 18-64 |  |  |  | Yes | Yes |
|  | 65+ |  |  |  | Yes | Yes |
| Unusual site thrombosis with<br>thrombocytopenia | 6 mos.-4/6 mos.-5 | [Definition below]** | 365 | 1-28<sup>50</sup> | Yes | No |
|  | 5-17/6-17 |  |  |  | Yes | No |
|  | 18-64 |  |  |  | Yes | Yes |
|  | 65+ |  |  |  | Yes | Yes |

While most outcomes underwent sequential testing, outcomes were only descriptively monitored if there were limited historical case counts to estimate background rates, which were required for testing. Among children aged 6 months – 4 years and 6 months – 5 years receiving BNT162b2 and mRNA-1273.222 vaccines respectively, 12 outcomes underwent descriptive monitoring and sequential testing, while eight were only descriptively monitored. For persons 5-17 years and 6-17 years receiving BNT162b2 and mRNA-1273.222 vaccines respectively, 13 outcomes were both descriptively monitored and sequentially tested, and eight outcomes were only descriptively monitored. Seventeen outcomes were included in descriptive monitoring and sequential testing for adults 18 years and older, while a single outcome was only descriptively monitored (Table 4).

#### 2.5 Statistical Analyses

##### Descriptive Monitoring

Vaccine uptake and health outcome counts were monitored monthly along with the demographic characteristics of vaccine recipients. The frequency of concomitant bivalent COVID-19 and seasonal influenza vaccination was also monitored. Prevalence of concomitant influenza vaccination was measured on the same-day and within 42 days prior to or following the bivalent COVID-19 mRNA vaccination date. Concomitant influenza vaccination was only descriptively monitored and was not included in inferential testing nor adjusted for in any analyses.

##### Sequential Testing

The Poisson Maximized Sequential Probability Ratio Test (PMaxSPRT) was used to evaluate the rate of outcomes in specific age groups following vaccination compared to a historical baseline rate.^6^ Monthly sequential testing was performed to generate incidence rate ratios (IRRs) of observed outcome rates compared with database-specific historical (expected) rates. Historical rates were adjusted to account for claims processing delay, and where case counts permitted, standardized by age and sex for the population younger than 65 years old, and by age, sex, race, and nursing home residency for the population 65 years and older.^7^ We estimated annual historical rates for a pre-COVID-19 period (2017-2019) and a COVID-19 period between April and December 2020. Selection of the comparator rate was based on the overlap between the 95% confidence intervals for the periods. If rates in the historical period (individual years between 2017 and 2020) did not have overlapping 95% confidence intervals indicating a substantive difference in the rates, we selected the lower or more stable rate as the most conservative approach. This was performed to enhance the sensitivity of the test by increasing the likelihood that potential signals were not missed. Otherwise, the median annual rate was selected. Pre-COVID-19 rates from 2019 were selected for most outcomes; however, rates from the 2020 COVID-19 period were selected if the COVID-19 period rates did not return to pre-COVID-19 levels.

For each outcome, monthly sequential testing was conducted in each database and began when a minimum of three outcomes after bivalent COVID-19 vaccination was observed. Testing was conducted until the earlier of one of the following events occurred: an observed signal, surveillance end, or when the outcome-specific prespecified surveillance length was met, which was defined as the expected number of outcomes within a 6-month period.^5^ We used a one-tailed test with a null hypothesis that the observed outcome rate was no greater than the historical comparator beyond a prespecified test margin with an overall alpha of 1%. A strict alpha level was selected to minimize false signals based on the large number of tests performed. Testing margins were determined for each outcome based on input from clinicians to avoid detection of minimal increases in risk that were unlikely to be clinically relevant. A statistical signal occurred if the log likelihood ratio exceeded the critical value, which was a threshold set to determine if the observed result was due to chance.^5^ SAS © version 9.4, R version 4.1.2, and R Sequential Package Version 3.3.1 were used for the analysis.

This surveillance activity was performed under the FDA Biologics Effectiveness and Safety (BEST) Initiative and is covered under the scope of the FDA public health surveillance mandate.

### 3. Results

#### Descriptive Monitoring

A total of 13.9 million bivalent COVID-19 mRNA vaccine doses were observed, including 5.5 million vaccinations across commercial databases including Carelon Research, CVS Health, and Optum databases, and 8.4 million vaccinations in the Medicare FFS database (Table 2). Figure 1 shows weekly vaccine uptake by vaccine brand and database. Among commercial databases, Carelon had the highest weekly vaccine uptake compared to CVS Health and Optum databases. Figure 2 shows weekly vaccine uptake in commercial and Medicare databases by vaccine brand and age group. For adults 18 years and older, peak vaccination uptake occurred between September and December 2022, with the highest weekly vaccination uptake observed for persons 65 years and older. For children 5-17 years, BNT162b2 vaccine recipients had the highest weekly vaccine uptake between September and December 2022. Minimal weekly vaccine uptake was observed for BNT162b2 and mRNA-1273.222 vaccine users 6 months-4 years and 6 months-5 years respectively, and mRNA-1273.222 vaccine users 6-17 years.

**Figure 1.**
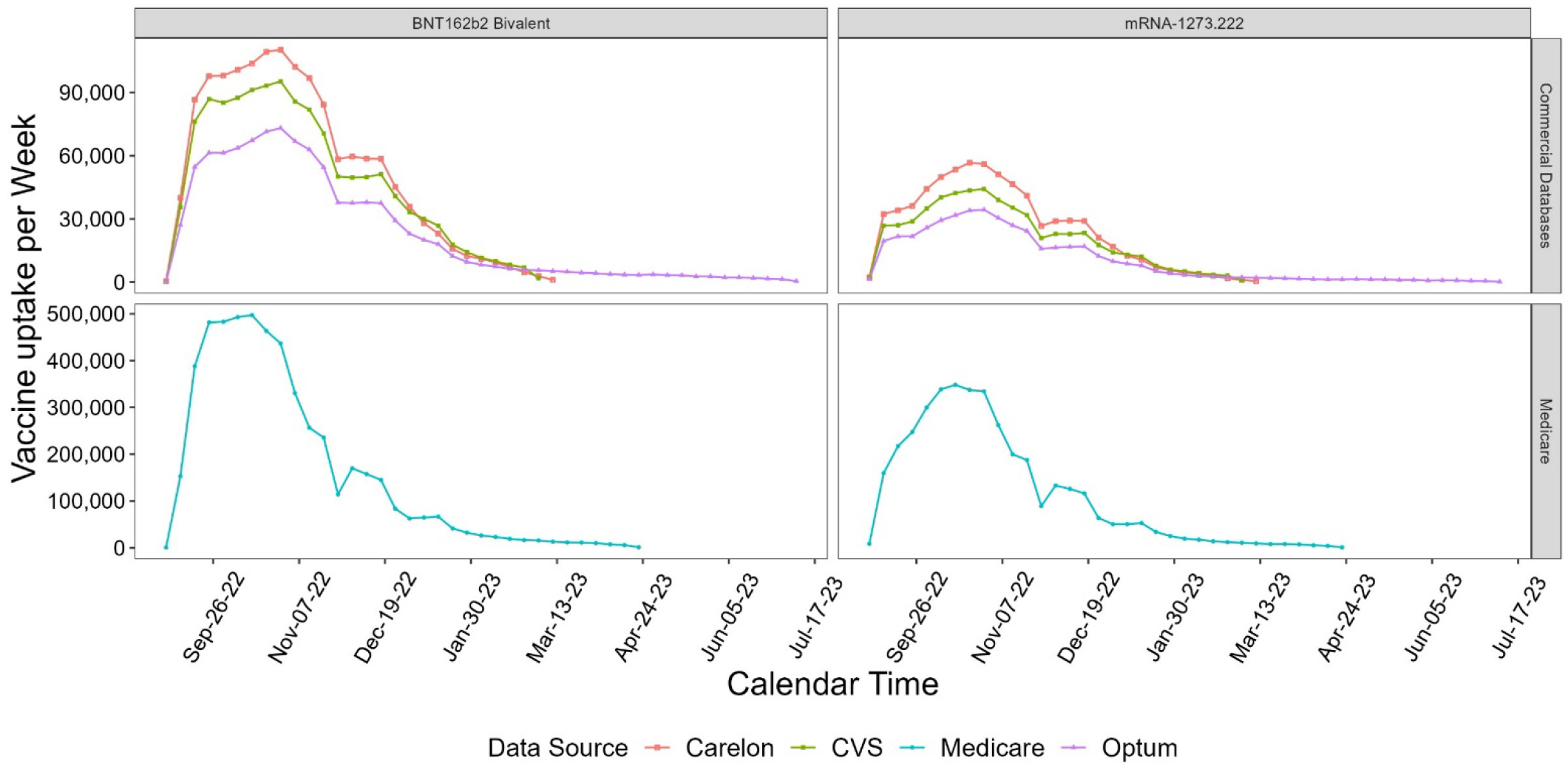
Pattern of vaccination trends in vaccine administrations of bivalent COVID-19 mRNA vaccines, August 2022 - April 2023 by vaccine brand and database.

**Figure 2.**
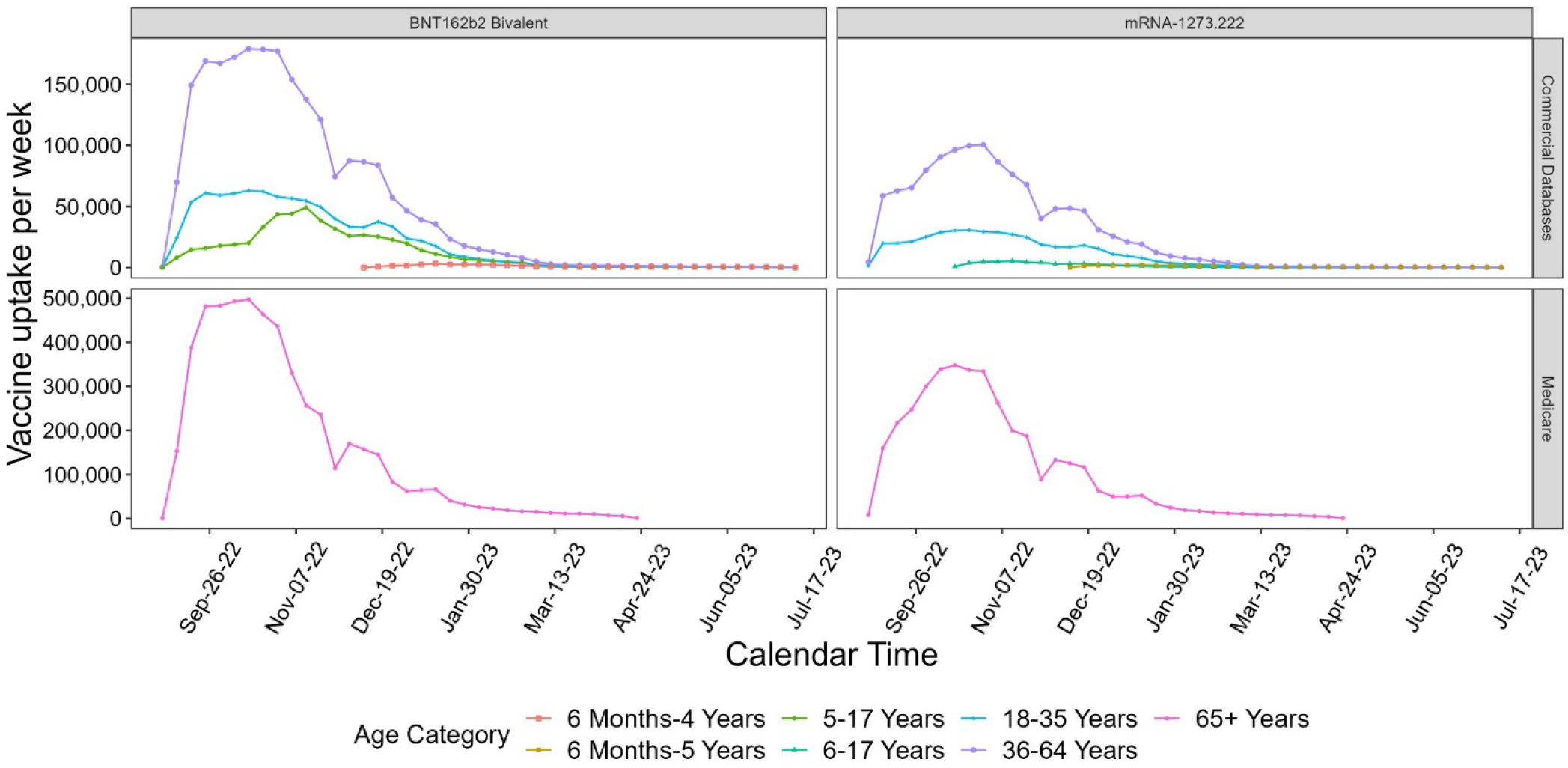
Pattern of bivalent COVID-19 mRNA vaccines uptake August 2022-July 2023 by age group and database.

**Table 2.** Characteristics of the population vaccinated with bivalent COVID-19 mRNA vaccines aged 6 months-64 years in CVS Health, Carelon Research, and Optum Databases; aged≥ 65+ years in Medicare Database.

| Patient Characteristics | Any Bivalent COVID-19 mRNA Vaccine |  |  |  | BNT162b2 Bivalent |  |  |  | mRNA-1273.222 |  |  |  |
| --- | --- | --- | --- | --- | --- | --- | --- | --- | --- | --- | --- | --- |
|  | Commercial Databases |  | Medicare |  | Commercial Databases |  | Medicare |  | Commercial Databases |  | Medicare |  |
|  | N | % | N | % | N | % | N | % | N | % | N | % |
| **Total** | **5,498,556** | **100** | **8,380,283** | **100** | **3,769,301** | **100** | **4,869,360** | **100** | **1,729,255** | **100** | **3,510,923** | **100** |
| **Age at Vaccine Administration (years)** |  |  |  |  |  |  |  |  |  |  |  |  |
| 6 mos.-4 | 33,051 | 0.6 | -- | -- | 33,051 | 0.9 | -- | -- | -- | -- | -- | -- |
| 5-11 | 234,201 | 4.3 | -- | -- | 234,201 | 6.2 | -- | -- | -- | -- | -- | -- |
| 6 mos.-5 | 23,864 | 0.4 | -- | -- | -- | -- | -- | -- | 23,864 | 1.4 | -- | -- |
| 6-11 | 21,719 | 0.4 | -- | -- | -- | -- | -- | -- | 21,719 | 1.3 | -- | -- |
| 12-17 | 329,297 | 6.0 | -- | -- | 301,069 | 8.0 | -- | -- | 28,228 | 1.6 | -- | -- |
| 18-25 | 454,374 | 8.3 | -- | -- | 319,187 | 8.5 | -- | -- | 135,187 | 7.8 | -- | -- |
| 26-35 | 872,613 | 15.9 | -- | -- | 579,921 | 15.4 | -- | -- | 292,692 | 16.9 | -- | -- |
| 36-45 | 1,026,398 | 18.7 | -- | -- | 684,872 | 18.2 | -- | -- | 341,526 | 19.7 | -- | -- |
| 46-55 | 1,128,330 | 20.5 | -- | -- | 748,022 | 19.8 | -- | -- | 380,308 | 22.0 | -- | -- |
| 56-64 | 1,374,709 | 25.0 | -- | -- | 868,978 | 23.1 | -- | -- | 505,731 | 29.2 | -- | -- |
| 65-74 | -- | -- | 4,414,887 | 52.7 | -- | -- | 2,544,098 | 52.2 | -- | -- | 1,870,789 | 53.3 |
| 75-84 | -- | -- | 2,937,151 | 35.0 | -- | -- | 1,699,813 | 34.9 | -- | -- | 1,237,338 | 35.2 |
| 85+ | -- | -- | 1,028,245 | 12.3 | -- | -- | 625,449 | 12.8 | -- | -- | 402,796 | 11.5 |
| Missing/<br>Unknown | 0 | 0.0 | 0 | 0.00 | 0 | 0.0 | 0 | 0.0 | 0 | 0.0 | 0 | 0.0 |
| **Sex** |  |  |  |  |  |  |  |  |  |  |  |  |
| Female | 2,973,454 | 54.1 | 4,760,468 | 56.8 | 2,039,091 | 54.1 | 2,784,173 | 57.2 | 934,363 | 54.0 | 1,976,295 | 56.3 |
| Male | 2,522,429 | 45.9 | 3,619,815 | 43.2 | 1,728,476 | 45.9 | 2,085,187 | 42.8 | 793,953 | 45.9 | 1,534,628 | 43.7 |
| Missing/<br>Unknown | 2,673 | 0.0 | 0 | 0.0 | 1,734 | 0.0 | 0 | 0.0 | 939 | 0.1 | 0 | 0.0 |
| **Urban/Rural** |  |  |  |  |  |  |  |  |  |  |  |  |
| Rural | 270,566 | 4.9 | 1,437,843 | 17.2 | 155,304 | 4.1 | 706,057 | 14.5 | 115,262 | 6.7 | 731,786 | 20.8 |
| Urban | 5,224,680 | 95.0 | 6,906,602 | 82.4 | 3,611,592 | 95.8 | 4,143,587 | 85.1 | 1,613,088 | 93.3 | 2,763,015 | 78.7 |
| Missing/<br>Unknown | 3,310 | 0.1 | 35,838 | 0.4 | 2,405 | 0.1 | 19,716 | 0.4 | 905 | 0.1 | 16,122 | 0.5 |
| **Nursing Home**<br>**Residency** |  |  |  |  |  |  |  |  |  |  |  |  |
| Nursing Home | -- | -- | 162,676 | 1.9 | -- | -- | 114,823 | 2.4 | -- | -- | 47,853 | 1.4 |
| Non-Nursing Home | -- | -- | 8,217,607 | 98.1 | -- | -- | 4,754,537 | 97.6 | -- | -- | 3,463,070 | 98.6 |
| **HHS Region\*\*** |  |  |  |  |  |  |  |  |  |  |  |  |
| Region 1 | 415,184 | 7.6 | 636,543 | 7.6 | 260,059 | 6.9 | 375,493 | 7.7 | 155,125 | 9.0 | 261,050 | 7.4 |
| Region 2 | 717,615 | 13.1 | 772,359 | 9.2 | 497,768 | 13.2 | 445,935 | 9.2 | 219,847 | 12.7 | 326,424 | 9.3 |
| Region 3 | 663,777 | 12.1 | 1,041,119 | 12.4 | 456,144 | 12.1 | 605,595 | 12.4 | 207,633 | 12.0 | 435,524 | 12.4 |
| Region 4 | 700,716 | 12.7 | 1,436,145 | 17.1 | 484,902 | 12.9 | 780,141 | 16.0 | 215,814 | 12.5 | 656,004 | 18.7 |
| Region 5 | 840,248 | 15.3 | 1,510,948 | 18.0 | 594,476 | 15.8 | 933,502 | 19.2 | 245,772 | 14.2 | 577,446 | 16.4 |
| Region 6 | 335,791 | 6.1 | 712,875 | 8.5 | 222,060 | 5.9 | 376,103 | 7.7 | 113,731 | 6.6 | 336,772 | 9.6 |
| Region 7 | 196,416 | 3.6 | 486,163 | 5.8 | 143,920 | 3.8 | 309,893 | 6.4 | 52,496 | 3.0 | 176,270 | 5.0 |
| Region 8 | 243,198 | 4.4 | 308,916 | 3.7 | 178,681 | 4.7 | 209,853 | 4.3 | 64,517 | 3.7 | 99,063 | 2.8 |
| Region 9 | 1,162,526 | 21.1 | 1,076,085 | 12.8 | 763,870 | 20.3 | 586,787 | 12.1 | 398,656 | 23.1 | 489,298 | 13.9 |
| Region 10 | 220,579 | 4.0 | 391,565 | 4.7 | 165,541 | 4.4 | 241,908 | 5.0 | 55,038 | 3.2 | 149,657 | 4.3 |
| Missing/<br>Unknown | 2,506 | 0.0 | 7,565 | 0.1 | 1,880 | 0.0 | 4,150 | 0.1 | 626 | 0.0 | 3,415 | 0.1 |
| **Concomitant Vaccination Status** |  |  |  |  |  |  |  |  |  |  |  |  |
| Influenza (Same<br>Day) | 2,223,077 | 40.4 | 2,764,112 | 33.0 | 1,545,503 | 41.0 | 1,720,039 | 35.3 | 677,574 | 39.2 | 1,044,073 | 29.7 |
| Influenza<br>(+/- 42 days)* | 3,391,431 | 61.7 | 6,005,972 | 71.7 | 2,325,523 | 61.7 | 3,539,611 | 72.7 | 1,065,908 | 61.6 | 2,466,361 | 70.2 |

\*Relative to bivalent COVID-19 mRNA vaccination date to align with broadest adverse event-specific risk interval
\*\*Health and Human Services (HHS) regions are designated by the U.S. HHS Department to serve state and local organizations. Information on individual U.S. states and territories included in each region is linked.
Data cuts: CVS Health (2/28), Carelon (3/4), Optum (7/8), Medicare (4/22)

Among vaccine recipients 6 months-64 years in commercial insurance databases, 12% were 6 months-17 years, 24% were 18-35 years, 39% were 36-55 years, and 25% were 56-64 years (Table 2). There was a slightly higher proportion of female recipients (54%) than males, which was consistent for BNT162b2 and mRNA-1273.222 vaccine brands. The majority of vaccine users resided in urban areas (95%). The prevalence of same-day concomitant bivalent COVID-19 and seasonal influenza vaccination was 40%, increasing to 62% in the 42 days prior to or after the COVID-19 vaccination date. There was a slightly higher proportion of same-day concomitant vaccination among BNT162b2 recipients (41%) than mRNA-1273.222 recipients (39%), increasing to 62% for both vaccine brands in the 42 days before or after bivalent COVID-19 vaccination (Table 2).

In the Medicare FFS population 65 years and older, 88% of vaccine recipients were aged 65-84 years with a higher proportion of females (57%) than males. This was relatively consistent across BNT162b2 and mRNA-1273.222 vaccine users (Table 2). The majority of COVID-19 vaccine users resided in urban areas (82%), with a slightly higher proportion of BNT162b2 vaccine users residing in urban areas (85%) compared to mRNA-1273.222 users (79%). Only a small portion of vaccine users resided in the nursing home (2%). The prevalence of same-day concomitant bivalent COVID-19 and seasonal influenza vaccination was 33%, increasing to 72% in the 42 days before or after bivalent COVID-19 vaccination. There was a higher rate of same-day concomitant vaccination with influenza vaccines among BNT162b2 vaccine users (35%) compared to mRNA-1273.222 users (30%), increasing to 73% and 70% respectively in the 42 days prior to or following COVID-19 vaccination.

#### Sequential Testing

Sequential testing was performed for outcomes with estimable background rates. Of 17 outcomes monitored with sequential testing in adults aged 18-64 years, anaphylaxis and myocarditis/pericarditis met the statistical threshold for a signal in one of three databases (Table 3). An anaphylaxis signal was detected following bivalent BNT162b2 and mRNA-1273.222 vaccination among participants aged 18-64 years in the Carelon Research database, based on <11 events^1^ for each vaccine brand. Similarly, a myocarditis/pericarditis signal was observed among bivalent BNT162b2 vaccine recipients aged 18-35 years in the Carelon Research database, based on <11 events^1^. No statistical signals were observed in CVS Health, Optum, or Medicare FFS databases.

**Table 3.**
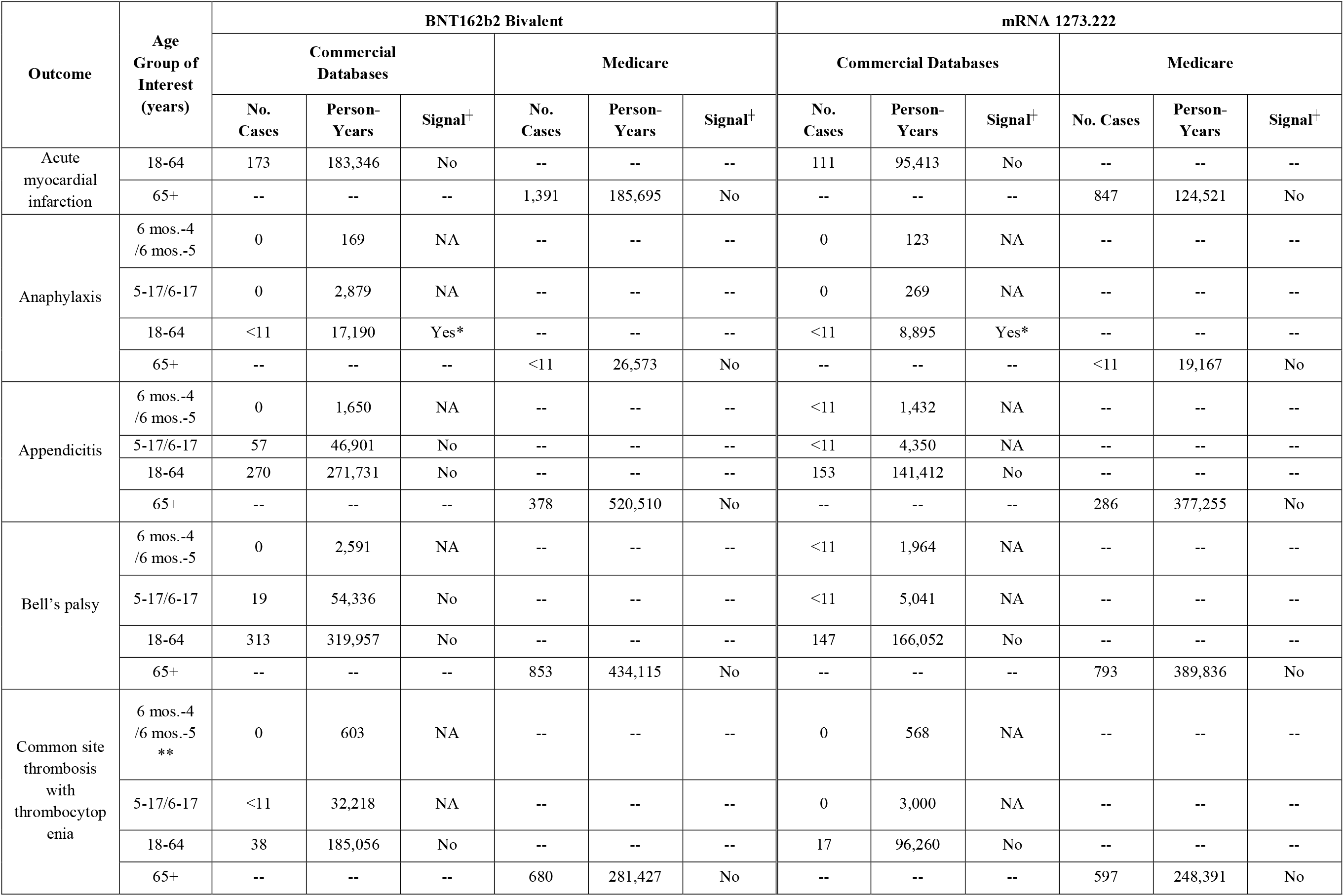
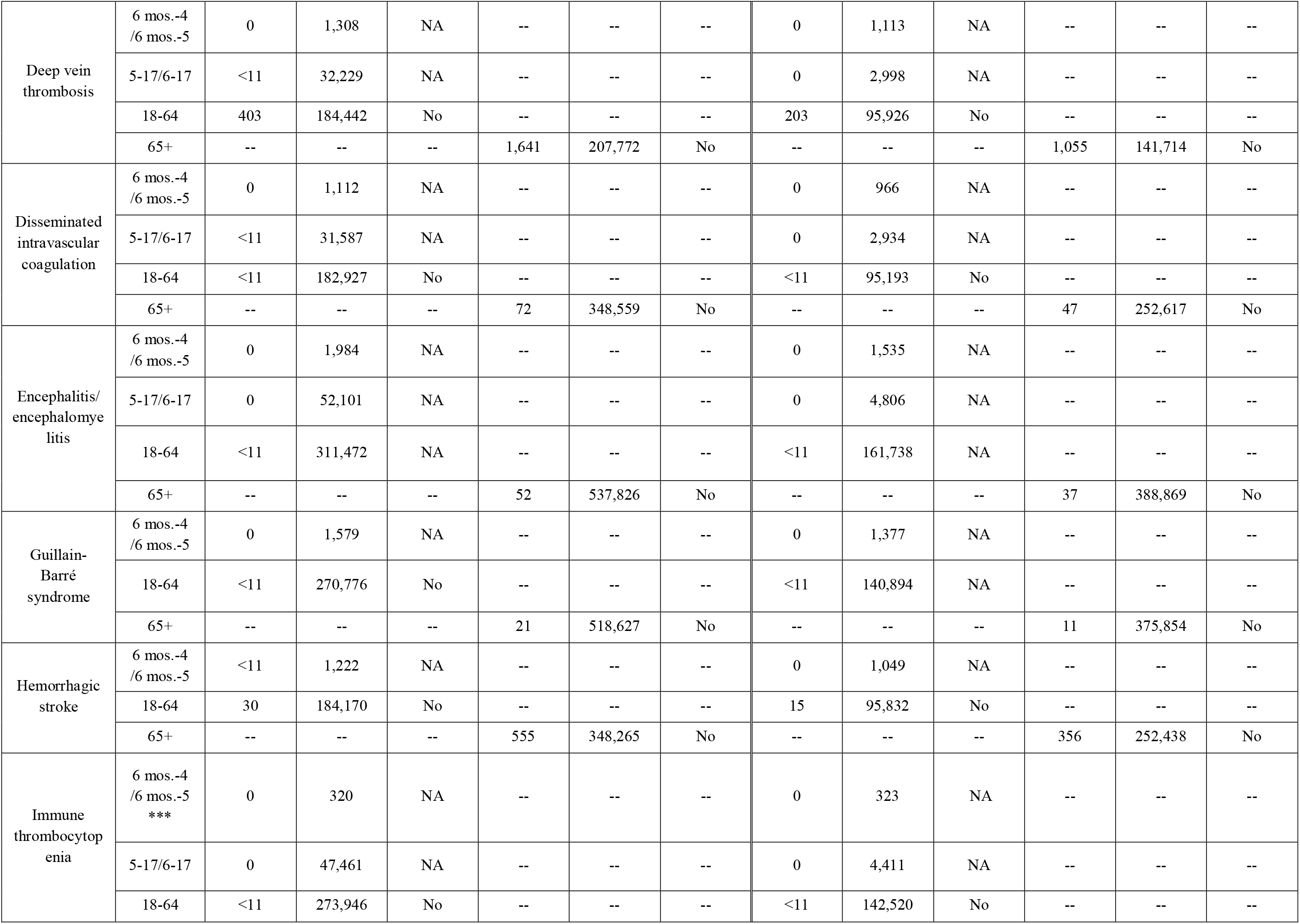
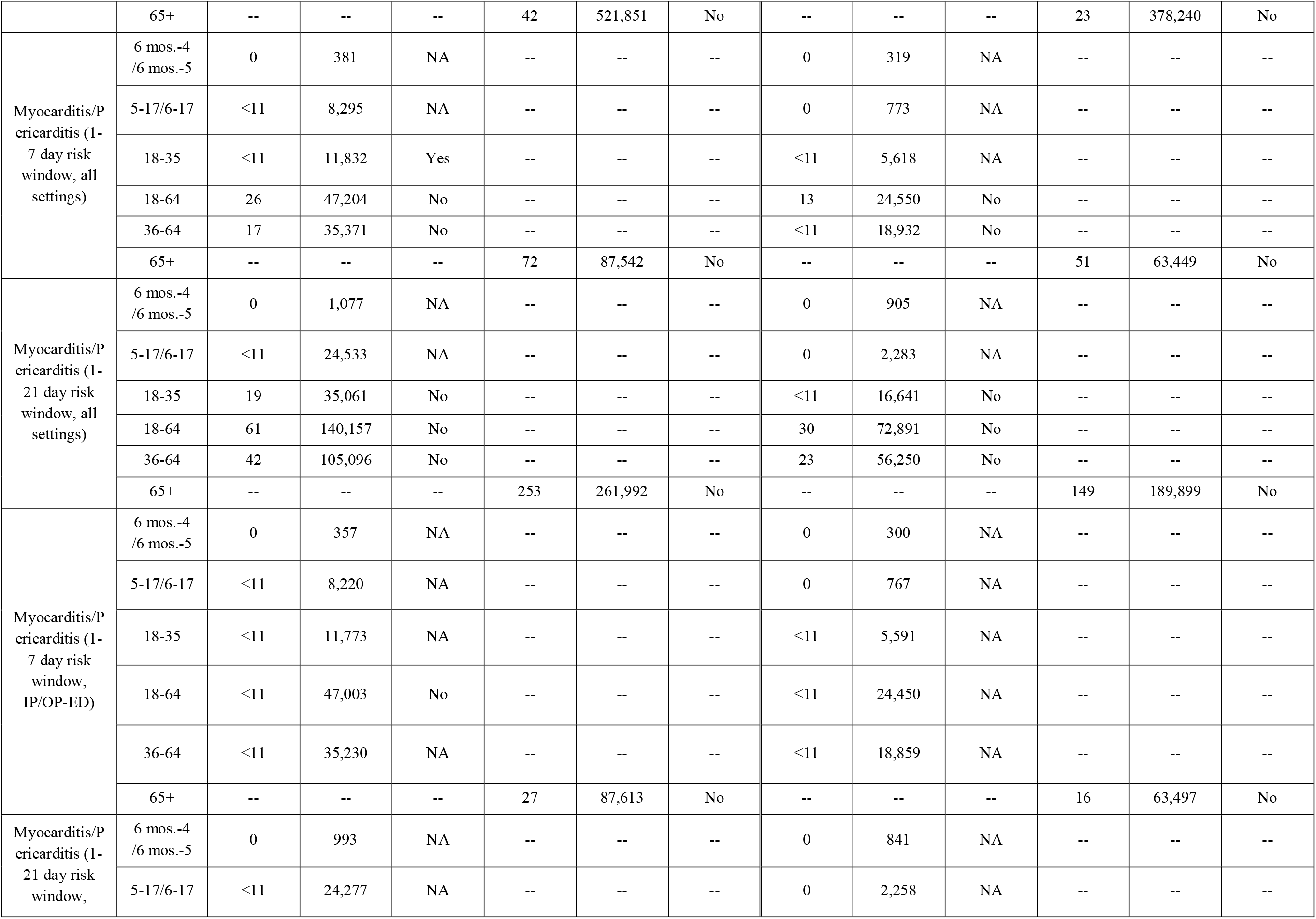
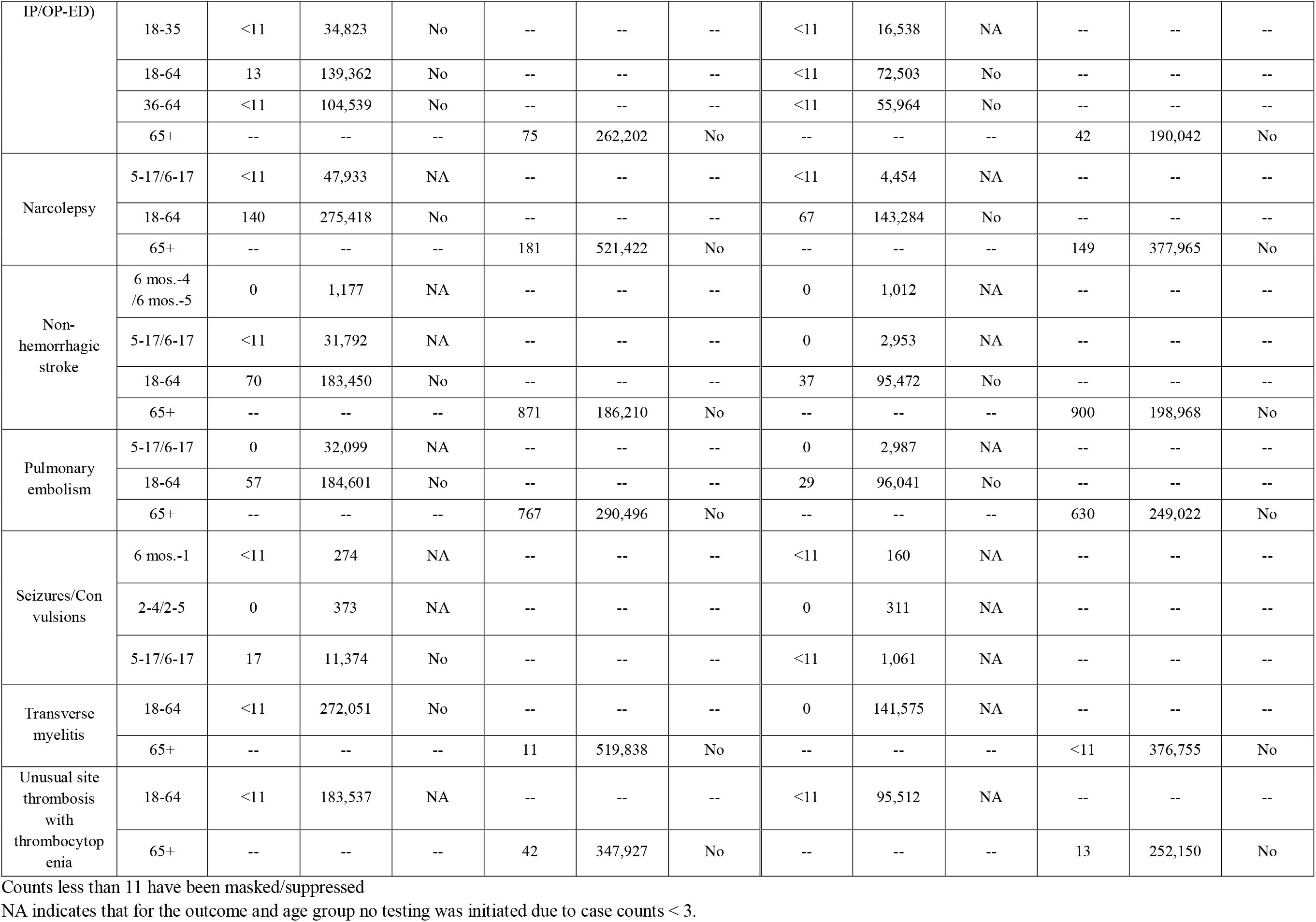
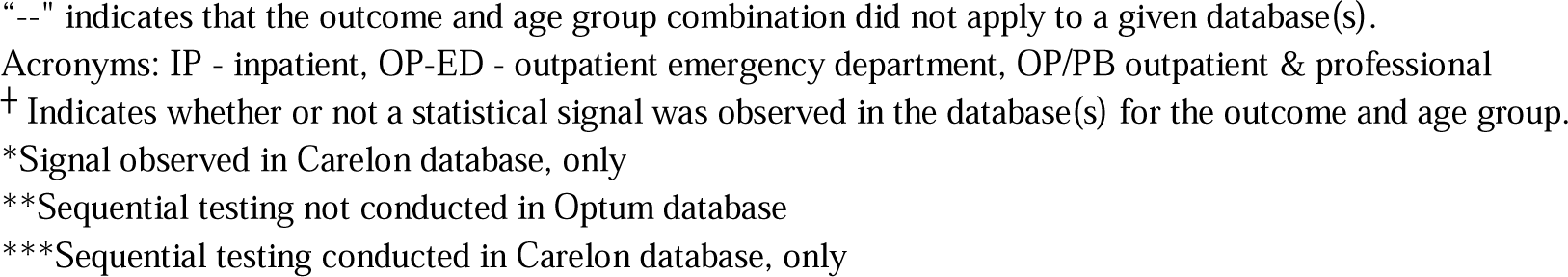
Sequential testing results in population vaccinated with bivalent COVID-19 mRNA vaccines.

**Table 4.** Frequency of vaccine doses and outcomes among outcomes only descriptively monitored.

| Outcome | Age Group of Interest (years) | BNT162b2 Bivalent |  |  |  | mRNA 1273.222 |  |  |  |
| --- | --- | --- | --- | --- | --- | --- | --- | --- | --- |
|  |  | Commercial Databases |  | Medicare |  | Commercial Databases |  | Medicare |  |
|  |  | No. Cases | No. Doses | No. Cases | No. Doses | No. Cases | No. Doses | No. Cases | No. Doses |
| Acute myocardial infarction | 6 mos.-4 / 6 mos.-5 | 0 | 20,857 | -- | -- | 0 | 17,361 | -- | -- |
|  | 5-17/6-17 | 0 | 436,784 | -- | -- | 0 | 40,746 | -- | -- |
| Common site thrombosis with thrombocytopenia | 6 mos.-4 / 6 mos.-5 * | 0 | 9,764 | -- | -- | 0 | 7,551 | -- | -- |
| Febrile seizures | 6 mos.-4 / 6 mos.-5 | <11 | 31,571 | -- | -- | <11 | 22,990 | -- | -- |
|  | 5-17/6-17 | <11 | 525,424 | -- | -- | <11 | 49,056 | -- | -- |
| Guillain-Barré syndrome | 5-17/6-17 | 0 | 436,785 | -- | -- | 0 | 40,746 | -- | -- |
| Hemorrhagic stroke | 5-17/6-17 | <11 | 436,775 | -- | -- | 0 | 40,746 | -- | -- |
| Immune thrombocytopenia | 6 mos.-4 / 6 mos.-5 ** | 0 | 15,111 | -- | -- | 0 | 12,432 | -- | -- |
| Kawasaki disease | 6 mos.-4 / 6 mos.-5 | 0 | 20,837 | -- | -- | 0 | 17,348 | -- | -- |
|  | 5-17/6-17 | <11 | 436,688 | -- | -- | <11 | 40,736 | -- | -- |
| Multisystem inflammatory syndrome | 6 mos.-4 / 6 mos.-5 | 0 | 20,856 | -- | -- | 0 | 17,359 | -- | -- |
|  | 5-17/6-17 | 0 | 436,772 | -- | -- | 0 | 40,745 | -- | -- |
|  | 18-64 | 0 | 2,478,389 | -- | -- | 0 | 1,288,891 | -- | -- |
|  | 65+ | -- | -- | <11 | 4,574,507 | -- | -- | 0 | 3,315,135 |
| Narcolepsy | 6 mos.-4 / 6 mos.-5 | 0 | 20,856 | -- | -- | 0 | 17,361 | -- | -- |
| Pulmonary embolism | 6 mos.-4 / 6 mos.-5 | 0 | 20,857 | -- | -- | 0 | 17,361 | -- | -- |
| Transverse myelitis | 6 mos.-4 / 6 mos.-5 | 0 | 20,856 | -- | -- | 0 | 17,361 | -- | -- |
|  | 5-17/6-17 | <11 | 436,784 | -- | -- | 0 | 40,746 | -- | -- |
| Unusual site thrombosis with thrombocytopenia | 6 mos.-4 / 6 mos.-5 | 0 | 20,857 | -- | -- | 0 | 17,361 | -- | -- |
|  | 5-17/6-17 | 0 | 436,784 | -- | -- | 0 | 40,746 | -- | -- |

Counts less than 11 have been masked/suppressed.
"--" indicates that the outcome and age group combination did not apply to a given database(s).
\*Contain counts from Optum, only
\*\*Contain counts from CVS Health and Optum, only
Data cuts: CVS Health (2/28), Carelon (3/4), Optum (7/8), Medicare (4/22)

The incidence rate of anaphylaxis following bivalent BNT162b2 and mRNA-1273.222 vaccination was 74.5 and 109.4 cases per 100,000 person-years respectively, among those 18-64 years in the Carelon database. The incidence rate of myocarditis/pericarditis following bivalent BNT162b2 vaccination was 131.4 cases per 100,000 person-years for persons 18-35 years in the Carelon database.

Among the population aged 6 months-4 years and 6 months-5 years receiving the bivalent COVID-19 BNT162b2 and mRNA-1273.222 vaccines, respectively, sequential testing was not initiated for any of the 12 outcomes for which testing was planned, as the minimum number of cases needed to start testing (i.e., 3 cases) was not observed. Similarly, for the population aged 5-17 years, of 13 outcomes included in testing, testing was only initiated for appendicitis, seizures/convulsions, and Bell’s Palsy following BNT162b2 vaccination across commercial databases due to limited case counts for the other outcomes. Testing was not initiated for any outcomes among mRNA-1273.222 vaccine recipients aged 5-17 years due to limited case counts. The majority of outcomes did not initiate testing in persons 6 months-17 years due to limited case counts. Of outcomes that initiated testing in this age group, no statistical signals were detected for either vaccine brand (Table 3).

In the Medicare population aged 65 years and older, sequential testing was initiated for all 17 outcomes for both BNT162b2 and mRNA-1273.222 vaccinations, and no statistical signals were detected (Table 3).

### 4. Discussion

This study monitored the safety of bivalent COVID-19 mRNA vaccines administered to 13.9 million persons 6 months and older in the U.S. by evaluating the risk of several outcomes following vaccination. Two outcomes met the statistical threshold for a signal in one of three commercial databases: anaphylaxis following bivalent BNT162b2 and mRNA-1273.222 vaccination in recipients aged 18-64 years and myocarditis/pericarditis following BNT162b2 vaccination in recipients aged 18-35 years.

These findings are largely consistent with existing safety assessments of COVID-19 mRNA vaccines. An elevated risk of myocarditis/pericarditis and anaphylaxis was identified following monovalent mRNA vaccines.^8–12^ Risk of myocarditis/pericarditis was specifically identified in males aged 12-39 years of age in other studies.^8,10,12^ While an increased anaphylaxis risk has been reported,^9,11,13^ more recent studies have suggested a comparable risk to that of other vaccines.^14^ Surveillance for the bivalent COVID-19 mRNA vaccines by the Vaccine Adverse Event Reporting System (VAERS) has similarly shown cases of myocarditis/pericarditis and allergic/anaphylactic events among the population aged 12 years and older.^15^ A prior near-real time surveillance study identified a statistical signal for seizures/convulsions following monovalent BNT162b2 and mRNA-1273 vaccination in children 2-4/5 years.^16^ Our study identified zero cases of seizures/convulsions in this age group following bivalent vaccination, and no outcomes met the statistical threshold for signal in the young age group.

Our study population had a high prevalence of same-day concomitant administration of bivalent COVID-19 mRNA and influenza vaccines. Clinical trials that have assessed the safety and reactogenicity of concomitant administration of these vaccines have not found any safety concerns or reduced immune response related to the concomitant use of these vaccines.^17–19^ While we had a high prevalence of concomitant vaccination, our study did not perform testing stratified by concomitant influenza vaccination status; thus, we are unable to draw any conclusions about the safety of concomitant vaccination.

This study had several strengths. The study included a large sample size of geographically diverse participants from all age groups authorized to receive bivalent BNT162b2 or mRNA-1273.222 vaccines from both commercially and publicly insured populations across the U.S. The study also evaluated several outcomes as vaccines were administered. The near-real time surveillance method allowed for vaccine safety to be monitored shortly after bivalent COVID-19 mRNA vaccine authorizations. Outcomes were similarly able to be monitored for an extensive period varying from three to eleven months depending on database, vaccine brand and age group.

The near-real time surveillance method applied in this study is a crude signal detection method used for rapid safety screening. This method uses a comparator (i.e., aggregate historical rates) with limited adjustment for confounding factors. This method, similarly, requires the specification of multiple parameters that, if misspecified, could affect the presence or absence of detected signals such as risk interval timing and length, and the testing margin used to identify meaningful elevations in risk. Additionally, we did not adjust for multiple testing of outcomes which may have increased the likelihood of observing a false positive safety signal when risk is not truly elevated. Therefore, this method does not establish a causal association between the vaccines and outcomes. Furthermore, although we observed substantial uptake of bivalent COVID-19 mRNA vaccines, this was reduced relative to administration of monovalent COVID-19 mRNA vaccine doses. This could have decreased our power to detect statistical signals, particularly for rare outcomes, and may explain some differences in signals detected or not detected across databases. Our study was also limited by the use of administrative claims data, collected for billing purposes, but used for safety surveillance activities. Our medical record review has shown that certain outcomes are detected in claims data more accurately than others.^20,21^ Since we did not conduct medical record review nor adjust for outcome misclassification for outcomes evaluated in this study, the presence and extent of outcome misclassification is unknown and could apply to at least some of the outcomes included in surveillance. This can bias signal detection in either direction.

The surveillance of bivalent COVID-19 mRNA vaccines supports the safety of these vaccines and is consistent with findings from surveillance results of monovalent COVID-19 mRNA vaccines. This study contributes to regulatory decision-making for COVID-19 vaccines and public health and supports the conclusion that the benefits of vaccination outweigh the risks.

All authors attest they meet the ICMJE criteria for authorship.

## Supporting information

COVID-19 Bivalent Safety Monitoring Manuscript Supplementary Tables

## Data Availability

The data used in the present study is confidential and is not available for reference.

## Acknowledgements

We would like to thank Bowen Chen, Yue Wu, Vincent Varvaro, Kamran Kazemi, Olivia Zhang, Nimesh Shah, Samikshya Siwakoti, Anchi Lo, Jing Wang, Bing Lyu from Acumen LLC; Grace Yang, Sarah Sargen, Alexandra Stone, Wafa Tarazi, Megan Ketchell, Kathryn Federici, Amaka Ume, Emily Myers, Eli Wolter, Jackson Slaney, Bobby Smith, Lauren Peetluk, and Elizabeth Bell from Optum; Anne Marie Kline, Nancy B. Shaik, Ana M. Martinez-Baquero, Smita Bhatia, Vaibhav Sharma from CVS; Shiva Vojjala, Ramya Avula, Shiva Chaudhary, Shanthi P Sagare, Ramin Riahi, Brian Greenwald, and Grace Stockbower, Michael Goodman, Michael Bruhn, and Ruth Weed from Carelon/IQVIA.

## Footnotes

1 Cell counts < 11 have been suppressed in the manuscript text and tables.

2 Employee of CVS Health.

3 Employee of Elevance Health Incorporated.

4 Employee of IQVIA.

