## Supplementary material for "Safety Monitoring of Bivalent COVID-19 mRNA Vaccines Among Recipients 6 months and Older in the United States": COVID-19 Bivalent Safety Monitoring Manuscript Supplementary Tables

**Table E1. Database characteristics and enrollment size**

| **Data Source** | **Claims Type** | **Update frequency** | **Data Lag*** | **Population Enrolled** | **Number of Immunization Information Systems (IIS) Jurisdictions**** |
| --- | --- | --- | --- | --- | --- |
| CVS Health† | Fully Adjudicated | Monthly | Approximately 80% data completeness in 3-4 months for inpatient claims, 2-3 months for outpatient claims, and 1-2 months for professional claims |  | 24 |
|  |  |  |  | 0-4 years: > 1.1 million |  |
|  |  |  |  | 5-11 years: > 1.5 million |  |
|  |  |  |  | 12-17 years: > 1.5 million |  |
|  |  |  |  | 18-64 years: > 14.5 million |  |
| Optum pre-adjudicated claims† | Pre-Adjudicated |  | Approximately 80% data completeness in 1-2 months for inpatient, outpatient, and professional claims | 0-4 years: > 0.9 million | 22 |
|  |  | Bi-Weekly |  | 5-11 years: > 1.3 million |  |
|  |  |  |  | 12-17 years: > 1.2 million |  |
|  |  |  |  | 18-64 years: > 11.5 million |  |
| Carelon Research† | Fully adjudicated | Monthly | Approximately 80% data completeness in 2-3 months for inpatient claims and 1-2 months for outpatient and professional claims |  | 9 |
|  |  |  |  | 0-4 years: > 1.3 million |  |
|  |  |  |  | 5-11 years: > 1.8 million |  |
|  |  |  |  | 12-17 years: > 1.8 million |  |
|  |  |  |  | 18-64 years: > 16.9 million |  |
| CMS Medicare Shared Systems Fee-for-Service Data (SSD) | Pre-Adjudicated | Daily | >80% data completeness in 30-70 days for inpatient claims | >34 million beneficiaries annually | NA |

*Data lag is based on 2020 claims delay distribution

** For commercial insurers, local and state-based IIS data was linked to commercial claims databases to supplement the capture of patient’s
COVID-19 vaccination history. Once validated, vaccination information from select IIS jurisdictions is utilized to determine vaccination
status for members of the health database that can be matched to patients in the IIS database.

†Average number of annual enrollees in a given age category between 2018-2021

**Table E2. Dose-specific EUA dates for Bivalent COVID-19 vaccine products**

| **Brand** | **Age Group (years)** | **Assigned Dose-Group** | **Emergency Use Authorization Date** |
| --- | --- | --- | --- |
| BNT162b2 Bivalent | 12+ | Bivalent Booster | August 31, 2022 |
|  | 5-11 | Bivalent Booster | October 12, 2022 |
|  | 6 months-4 | Bivalent Primary Series Dose 3 | December 8, 2022 |
| mRNA-1273.222 | 18+ | Bivalent Booster | August 31, 2022 |
|  | 6-17 | Bivalent Booster | October 12, 2022 |
|  | 6 months-5 | Bivalent Booster | December 8, 2022 |
